# CGG expansion in *NOTCH2NLC* is associated with oculopharyngodistal myopathy with neurological manifestations

**DOI:** 10.1101/2020.10.16.20213785

**Authors:** Masashi Ogasawara, Aritoshi Iida, Theerawat Kumutpongpanich, Ayami Ozaki, Yasushi Oya, Hirofumi Konishi, Akinori Nakamura, Ryuta Abe, Hiroshi Takai, Ritsuko Hanajima, Hiroshi Doi, Fumiaki Tanaka, Hisayoshi Nakamura, Ikuya Nonaka, Zhaoxia Wang, Shinichiro Hayashi, Satoru Noguchi, Ichizo Nishino

**Author notes:** Corresponding author: Ichizo Nishino, MD, PhD, Department of Neuromuscular Research, National Institute of Neuroscience, National Center of Neurology and Psychiatry (NCNP), 4-1-1 Ogawahigashi, Kodaira, Tokyo 187-8502, Japan.

## Abstract

**Background:** Oculopharyngodistal myopathy (OPDM) is a rare hereditary muscle disease characterized by progressive distal limb weakness, ptosis, ophthalmoplegia, bulbar muscle weakness and rimmed vacuoles on muscle biopsy. Recently, CGG repeat expansions in the noncoding regions of two genes, *LRP12* and *GIPC1*, have been reported to be causative for OPDM. Furthermore, neuronal intranuclear inclusion disease (NIID) has been recently reported to be caused by CGG repeat expansions in *NOTCH2NLC*.

**Objectives:** To identify and to clinicopathologically characterize OPDM patients who have CGG repeat expansions in *NOTCH2NLC* (OPDM_NOTCH2NLC).

**Methods:** Two hundred eleven patients from 201 families, who were clinically or clinicopathologically diagnosed with OPDM or oculopharyngeal muscular dystrophy, were screened for CGG expansions in *NOTCH2NLC* by repeat primed-PCR. Clinical information and muscle pathology slides of the identified OPDM_NOTCH2NLC patients were re-reviewed. Intra-myonuclear inclusions were further evaluated by immunohistochemistry and electron microscopy.

**Results:** Seven Japanese OPDM patients had CGG repeat expansions in *NOTCH2NLC*. All seven patients clinically had ptosis, ophthalmoplegia, dysarthria, and muscle weakness, and myopathologically had intra-myonuclear inclusions stained with anti-poly-ubiquitinated proteins, anti-SUMO1 and anti-p62 antibodies, which are diagnostic of NIID typically on skin biopsy, in addition to rimmed vacuoles. Sample for electron microscopy was available only from one patient, which showed intranuclear inclusions of 12.6 ± 1.6 nm in diameter.

**Conclusions:** We identified seven OPDM_NOTCH2NLC patients. Our patients had various additional central and/or peripheral nervous system involvement, albeit all being clinicopathologically compatible; thus, diagnosed as having OPDM, expanding a phenotype of the neuromyodegenerative disease caused by CGG repeat expansions in *NOTCH2NLC*.

## INTRODUCTION

Oculopharyngodistal myopathy (OPDM) is a rare adult-onset hereditary muscle disease characterized clinically by progressive ocular, pharyngeal, and distal limb muscle involvement and pathologically by rimmed vacuoles in muscle fibers.^1 2^ Till date, two causative genes, *LRP12* and *GIPC1*, have been identified for OPDM (thereby, we refer to them as OPDM_LRP12 and OPDM_GIPC1, respectively), and in both diseases, CGG repeat expansions in the noncoding regions of the corresponding genes are believed to be the cause although the pathogenesis remains largely unknown.^3 4^ Recently, CGG repeat expansions in the noncoding region of *NOTCH2NLC* were reported as causative for neuronal intranuclear inclusion disease (NIID), a slowly progressive neurodegenerative disorder with eosinophilic intranuclear inclusions in the central and peripheral nervous systems and a variety of organs.^3 5 6^ Interestingly, certain NIID patients also manifest muscle weakness, dysarthria, and dysphagia, not fully but partially mimicking OPDM. Nevertheless, a diagnosis of OPDM has never been made in these patients most likely because ocular symptoms, an essential feature of OPDM, were lacking.^7 8^ On the other hand, rare OPDM patients were reported to accompany sensorineural hearing loss and demyelinating neuropathy.^9^ The presence of such NIID patients and OPDM patients raises a possibility that OPDM in certain patients, especially those with additional neurological manifestations, may be caused by the same pathogenic mechanism with NIID. Therefore, we evaluated CGG expansions in *NOTCH2NLC* in patients who were suspected to have OPDM.

## Materials and methods

### Inclusion criteria of the patients

The National Center of Neurology and Psychiatry (NCNP) functions as a referral center for muscle diseases in Japan, providing pathological and genetic diagnoses. Among the samples that were sent to the NCNP for diagnostic purposes from 1978 to 2020, we included 211 Japanese patients from 201 families who were clinically or clinicopathologically diagnosed with OPDM or oculopharyngeal muscular dystrophy (OPMD) based on a combination of at least two of ptosis, bulbar symptoms, or rimmed vacuoles on muscle biopsy but did not have the GCN repeat expansion in the polyadenylate binding protein nuclear 1 (*PABPN1*) gene, the causative factor for OPMD.^10^ All patients provided informed consent for the use of their samples for research after the diagnosis. This study was approved by the ethical committees of the NCNP (A2019-123).

### Clinical information

For patients with CGG expansions in *NOTCH2NLC*, clinical information including imaging data of muscle and brain provided by the physicians at the time of muscle biopsy and/or genetic analysis was reviewed.

### Genetic analysis

The 211 patients were screened for CGG expansions in *NOTCH2NLC* by repeat primed-PCR (RP-PCR), and the CGG repeat length in patients who had expanded CGG repeats was determined by fragment analysis and/or Southern blotting, as described previously.^3 5^ The CGG repeats in *LRP12* and *GIPC1* were also evaluated by RP-PCR, fragment analysis, and/or Southern blotting.^3 4^

### Muscle histology

We re-reviewed the muscle pathology slides of a battery of histochemical stains, which were prepared at the time of diagnosis. Immunohistochemical analysis of 8-µm-thick serial frozen sections was also performed using the following primary antibodies: anti-poly-ubiquitinated proteins (FK1, BML-PW8805, 1:100; Enzo Life Sciences), anti-SUMO-1 (D-11:sc-5308, 1:50; Santa Cruz Biotechnology), anti-phospho-p62/SQSTM1 (Ser351) (PM074, 1:500; Medical & Biological Laboratories), and anti-caveolin-3 (sc7665, 1:200; Santa Cruz Biotechnology). After incubation with primary antibodies, the sections were incubated with DAPI (Cellstain DAPI, 1:1000; Fujifilm) and Alexa Fluor 488-, 568-, and 647-conjugated secondary antibodies (1:600; Invitrogen). Images were taken by Keyence CCD camera (Keyence, Osaka, Japan). For the evaluation of regenerating fibers, we performed immunohistochemical analysis of serial frozen sections using neonatal myosin heavy chain (nMHC, NCL-MHCn, 1:20; Leica Biosystems Newcastle). We analyzed the frequency of regenerating fibers, fibers with internal nuclei, fibers with rimmed vacuoles, small angular fibers, and type 2C fibers in 200 randomly selected muscle fibers in each patient. We also analyzed the frequency of myonuclei positive with anti-p62, anti-poly-ubiquitinated protein, and anti-SUMO antibodies in 200 randomly selected myonuclei. Glutaraldehyde-fixed muscle sample for electron microscopy (EM) was available only from patient 2. On EM, we measured the diameter of randomly-chosen 75 intra-myonuclear filaments, which were straight, non-overlapped and well-demarcated.

### Skin histology

Skin biopsy sample was available from two patients (patients 1 and 7). Serial sections of 4-µm thickness were fixed in formalin and stained by hematoxylin and eosin (H&E) and anti-p62/SQSTM1 antibody (sc-28359, 1;200, Santa Cruz Biotechnology).

### Data availability

The data supporting the findings of this study are available from the corresponding author upon request.

## Results

We screened 211 patients with a clinical or clinicopathological diagnosis of OPDM or OPMD and identified seven unrelated patients with OPDM with CGG expansions in *NOTCH2NLC* (OPDM_NOTCH2NLC) by RP-PCR. Repeat units were determined by fragment analysis in five patients (patients 1, 4, 5, 6, and 7) and by Southern blotting in two patients (patients 2 and 3) (figure 1A–C). All patients had >100 repeat expansions, and patient 2 had two long CGG expansions, the longer one being approximately 674 repeats (figure 1A–C and table 1). None of the seven patients had expanded CGG repeats in *LRP12* and *GIPC1*.

**Table 1.** Clinical symptoms of patients with CGG expansion in *NOTCH2NLC*

| Patient ID | 1 | 2 | 3 | 4 | 5 | 6 | 7 |
| --- | --- | --- | --- | --- | --- | --- | --- |
| Sex | F | F | M | M | F | M | M |
| Onset age | Childhood | 20s | 10s | 40s | 50s | 60s | 20s |
| Repeat size | 139 | 217,674 | 83, 184 | 116 | 135 | 132 | 128 |
| Initial symptom | Dev | MW | MW, RPD | Ptois | RPD | MW | PD |
| Ptois | + | + | + | + | + | + | + |
| Ophthalmoplegia | + | + | + | + | + | + | + |
| Dysphagia | + | + | + | — | — | + | + |
| Dysarthria | + | + | + | + | + | + | + |
| Facial weakness | + | + | + | — | — | + | + |
| Limb muscle weakness | + | + | + | + | + | + | + |
| Distal/proximal/both | D (TA, Pe) | D > P | D > P | D (TA) > P | PU | D = P | D > P |
| Other muscle weakness | Neck | Neck | Neck | — | — | — | — |
| Muscle atrophy | Diffuse | Distal | Diffuse | Distal | — | — | Diffuse |
| DTR | ↓↓ | ↓↓ | ↓ | ↓ | ↓ | ↓↓ | ↓↓ |
| Sensory disturbance | — | — | — | + | + | + | — |
| Cerebellar symptoms Tremor | Ataxia | — | — | Tremor | Ataxia | NA | Ataxia |
| Eye abnormality | VD | RP | RPD | Mio, Pho | RPD | Cataract | — |
| Ear abnormality | Co | SHL | NA | + | + | + | — |
| HDSR | 20 | 30 | 29 | 28 | 6 | 20 | NA |
| Leukoencephalopathy | + | — | NA | NA | + | — | + |
| Neuropathy confirmed by NCV | + | NA | — | + | NA | + | + |
| ECG, UCG | — | Long QT | — | — | — | AV block | — |
| CSF cells per μl | 2 | NA | NA | 2 | 1 | 1 | 4 |
| CSF protein, mg/dl | 60 | NA | NA | 110 | 157 | 77 | 59 |
| CK IU/L | 203 | 654 | 1475 | 1886 | 63 | 735 | 436 |
| Lactate mg/dl | 19.6 | 14.0 | 17.3 | NA | 21.6 | 5.3 | 9.7 |

Co, conductive deafness; Dev, developmental delay; DL, distal lower limb; DTR, deep tendon reflex; DU, distal upper limb;
HDSR, Hasegawa dementia rating scale-revised; Mio, miosis; MW, muscle weakness; N, neck weakness; NA, not available;
Pe, peroneal; Pho, photophobia; PU, proximal upper limb; RP, retinal pigmentation; RPD, retinal pigmentary degeneration;
SHL, sensory hearing loss; TA, tibialis anterior; VD, visual disturbance;

**Figure 1.**
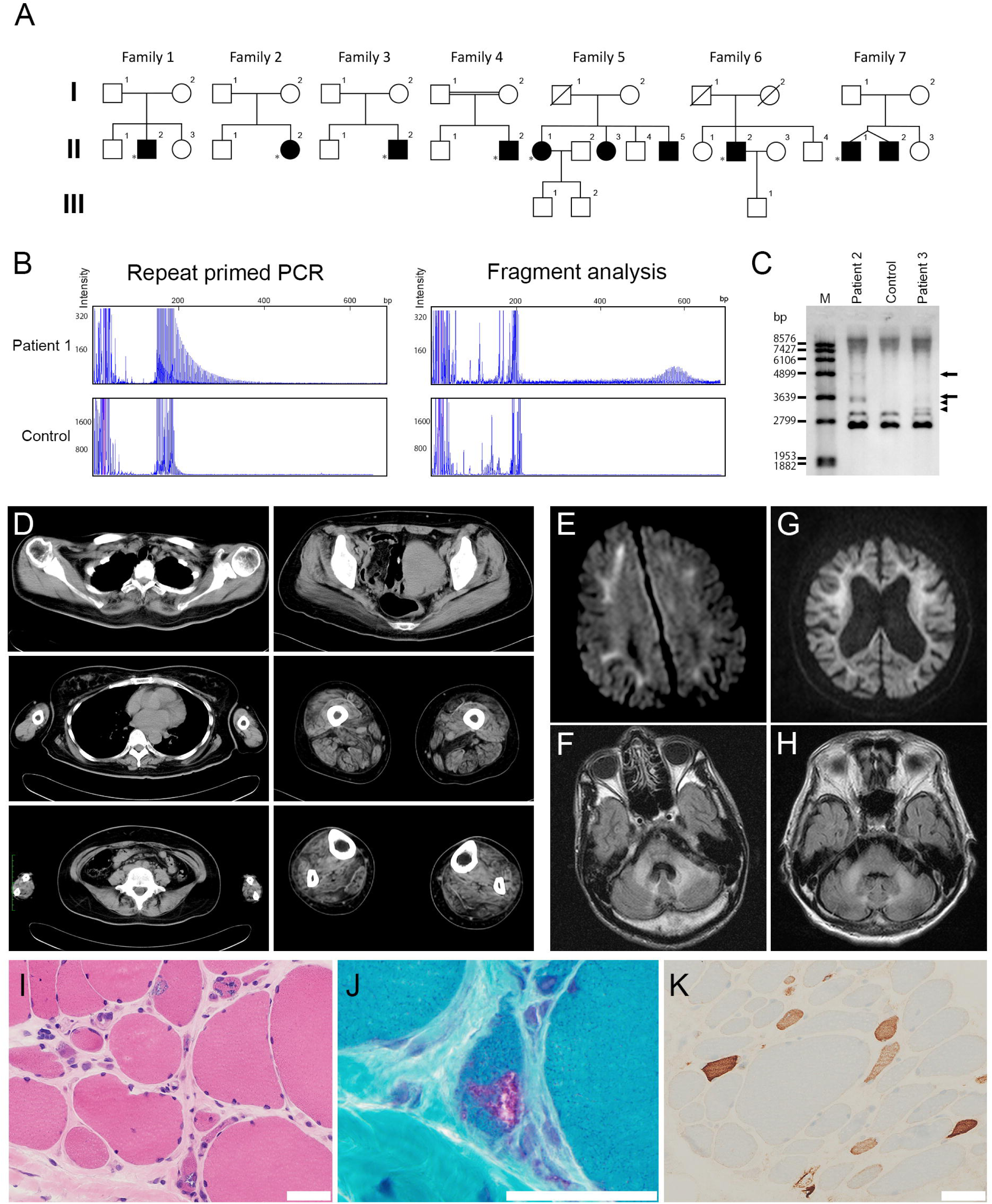
Clinical information of patients with CGG expansion in *NOTCH2NLC*. (A) Family trees of seven families. White box shows unaffected. Black box shows patients. * shows probands. (B) Repeat primed PCR and fragment analysis for patient 1 and control. (C) Southern blotting analysis for CGG expansion in *NOTCH2NLC*. The arrows and arrowheads indicate the expanded alleles in patients 2 and 3, respectively. M, DNA marker. (D) Imaging of muscle CT from patient 2 shows asymmetric muscle atrophy. Right gluteus maximus, left vastus lateralis, adductor magnus, and rectus femoris are severely affected. (E) DWI image of patient 1. (F) FLAIR image of patient 1. (G) DWI image of patient 5. (H) FLAIR image of patient 5. (I) Marked fiber size variation, moderate endomysial fibrosis, and fibers with internal nuclei are seen on H&E. (J) Rimmed vacuole is present in a muscle fiber on mGT. (K) Some myofibers express neonatal myosin heavy chain. (I–K) Scale bars denote 50 µm.

The clinical information of the seven patients with CGG expansions in *NOTCH2NLC* is summarized in table 1. The age at onset varied widely from infancy to 67 years. All the seven patients had ptosis, ophthalmoplegia, dysarthria, limb muscle weakness, and decreased deep tendon reflex. Dysphagia and facial muscle weakness were detected in five (71%) patients. Five (71%) patients had predominantly distal limb muscle weakness and muscle atrophy, whereas one (14%) patient had proximal upper limb muscle weakness. In addition to these clinical manifestations typical of OPDM, all patients had central or peripheral nervous system abnormalities such as leukoencephalopathy, retinal pigmentary degeneration, ataxia, tremor, deafness, peripheral neuropathy, and increased CSF protein levels. The CK level was moderately increased (436–1886 U/L) in five (71%) patients.

Muscle CT data of three patients were available. One of three patients had asymmetric muscle involvement with fat replacement markedly in the left adductor magnus and moderately in the right gluteus maximus, left vastus lateralis, and left rectus femoris (figure 1D). The other two patients showed muscle atrophy in calf muscles. Brain MR examinations were available in five patients (figure 1E–H). High-intensity signals were observed in the middle cerebellar peduncles on FLAIR images in three patients (patients 1, 5, and 7). Furthermore, high signals on FLAIR were observed in the medial part of the cerebellar hemisphere right beside the vermis and cerebral white matter in two patients (patients 1 and 5). High signals were noted along the corticomedullary junction on DWIs in two patients (patients 1 and 5). Moderate to marked ventricular enlargement was observed in three of five patients.

On muscle histology, all patients had fibers with rimmed vacuoles and small angular fibers (figure 1I,J, and table 2). The variation in fiber size was moderate to marked in six patients and mild in one patient. Six patients had regenerating fibers, whereas one patient had no necrotic or regenerating fibers (figure 1K and table 2). Only one patient (patient 3) had moderate endomysial fibrosis, whereas the other patients had no or only minimal endomysial fibrosis (figure 1I and table 2). Intra-myonuclear inclusions, primarily located in the center of the myonuclei, were detected by anti-poly-ubiquitinated protein, anti-SUMO1, and anti-phospho-p62/SQSTM1 antibodies in all patients (figure 2A–L). EM analysis of the muscle from patient 2 revealed tubulofilamentous inclusions without limiting membrane but with low-electron-density halo around the nucleus, located in the center of the myonuclei (figure 2M,N). The diameters of the 75 randomly-chosen filaments were 12.6 ± 1.6 nm (M ± SD).

**Table 2.** Pathological findings of patients with CGG expansion in *NOTCH2NLC*

| Patient ID | 1 | 2 | 3 | 4 | 5 | 6 | 7 |
| --- | --- | --- | --- | --- | --- | --- | --- |
| Sex | F | F | M | M | F | M | M |
| Age at muscle biopsy | 20s | 40s | 20s | 40s | 70s | 70s | 20s |
| Fiber size variation | Moderate | Marked | Marked | Mild | Moderate | Moderate | Moderate |
| Regenerating fibers | 2% | 2% | 16% | 1% | — | 0.5% | 5% |
| Small angular fibers | 4% | 10.5% | 5% | 1% | 8.5% | 8% | 10% |
| Endomysial fibrosis | ± | ± | ++ | ± | — | ± | ± |
| Fibers with internal nuclei | 2% | 5.5% | 4.5% | 19% | 1% | 17.5% | 3% |
| Fibers with rimmed vacuoles | 1.5% | 3.5% | 4% | 0.5% | 0.5% | 2% | 2.5% |
| Type 2C fibers | 1.5% | 1% | 2% | 1.5% | 1.5% | 2% | 3.5% |
| Intra-myonuclear p62 positive | 3.5% | 2.5% | 7% | 2.5% | 1% | 3% | 2% |
| Intra-myonuclear ubiquitin positive | 2% | 0.5% | 2.5% | 1.5% | 2.5% | 0.5% | 2.5% |
| Intra-myonuclear SUMO1 positive | 2.5% | 0.5% | 2.5% | 2.5% | 2.5% | 1% | 2% |

M: male; F: female

**Figure 2.**
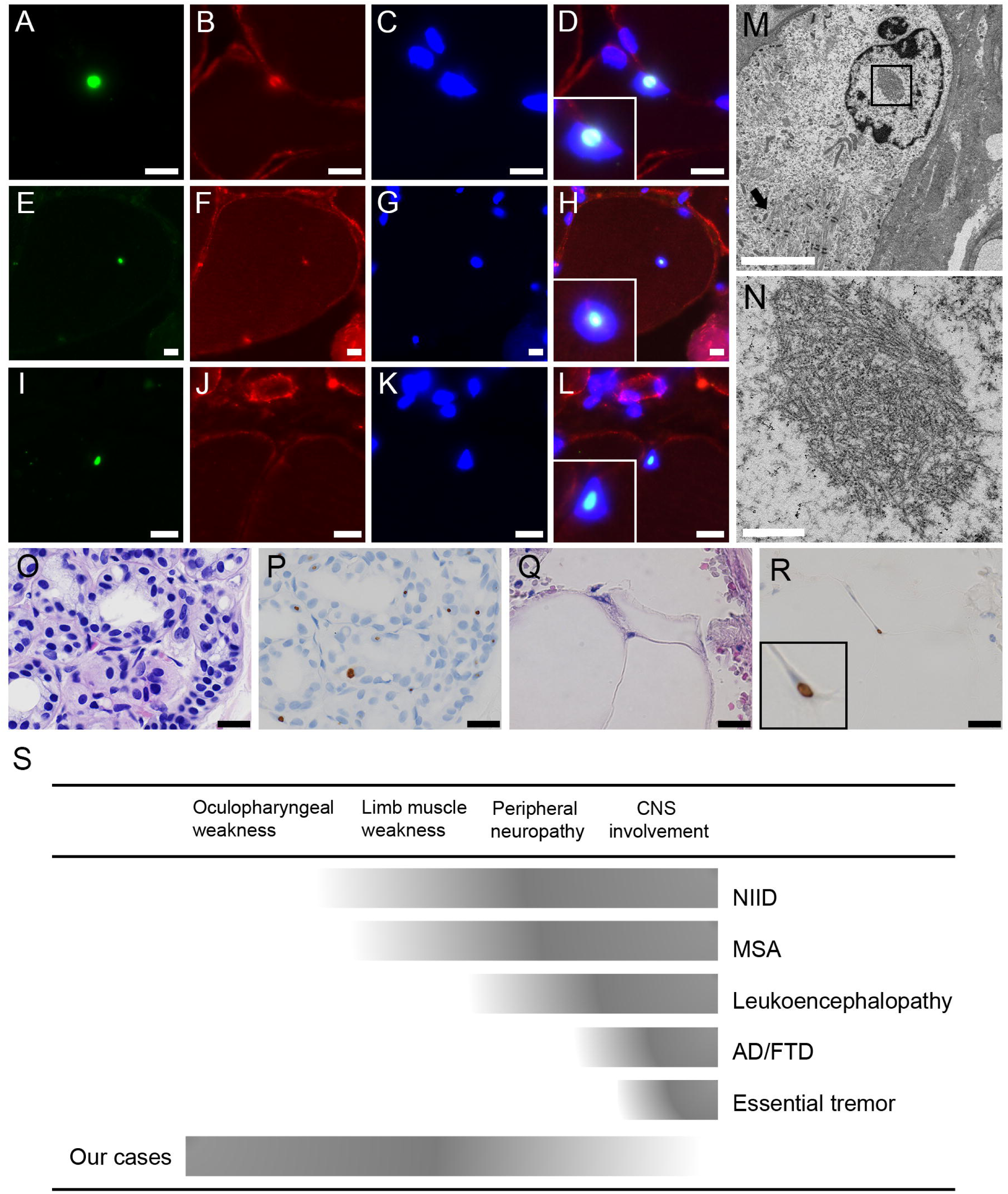
Pathology, revealed by electron microscopy and disease spectrum. Anti-poly-ubiquitinated protein antibody (A), anti-caveolin-3 antibody (B, F, J), DAPI (C, G, K), anti-SUMO-1 antibody (E), and anti-p62 (I) were stained. D, H, and L show merged immunohistochemistry. (A–L) Scale bars denote 10 µm. On electron microscopy, a longitudinal section shows tubulofilamentous inclusions within the myonuclei together with markedly disorganized myofibrils (arrow) in the surrounding area (M, N). (M) Scale bar denotes 5 µm. (N) Scale bar shows 500 nm. (O, Q) HE, (P, R) p62 were stained on serial sections of the skin sample in patient 1. Intranuclear inclusions with p62-positive in sweat gland cells (P) and adipocytes (R) are seen. (O–R) Scale bars denote 20 µm. (S) Disease spectrum caused by CGG expansion in *NOTCH2NLC*.

Skin biopsies from patients 1 and 7 showed p62-positive intranuclear inclusions (figure 2O–R).

## Discussion

NIID is a slowly progressive neurodegenerative disorder that is pathologically characterized by eosinophilic hyaline intranuclear inclusions in the central and peripheral nervous systems, as well as in the visceral organs and the skin. This disorder has been considered to be a heterogeneous disease because of the highly variable clinical manifestations.^7^ Recent studies have reported noncoding CGG repeat expansions in *NOTCH2NLC* as the causative factor for NIID.^3 5 6^ Subsequently, a variety of diseases have been associated with CGG repeat expansions in *NOTCH2NLC*, including multiple system atrophy (MSA), leukoencephalopathy, Alzheimer’s disease and frontotemporal dementia (AD/FTD), tremor, and retinal dystrophy, suggesting that the spectrum of *NOTCH2NLC* diseases is in fact wide (figure 2S).^11-15^

Interestingly, some NIID patients show additional myopathic features, such as limb muscle weakness, dysphagia, and dysarthria.^7 8^ However, neither ptosis/ophthalmoplegia, a clinical hallmark of OPDM, nor rimmed vacuole, a pathological hallmark of OPDM, has never been described in any histologically or genetically-confirmed NIID patient.^7 16^ Not surprisingly, to the best of our knowledge, none of the NIID patients has ever been clinicopathologically diagnosed with OPDM, and vice versa.

Although our OPDM_NOTCH2NLC patients were clinicopathologically diagnosed with OPDM, they had additional clinical manifestations that are partially reminiscent of NIID, including leukoencephalopathy and retinal degeneration. In addition, one and three patients respectively had tremor and ataxia. The MRI findings in two of five patients revealed high intensity signals in the middle cerebellar peduncles and in the paravermal area on FLAIR images and in the corticomedullary junction on DWI images,similarly to those in patients with NIID (figure 1E–H). ^3 17 18^

The identification of patients with OPDM_NOTCH2NLC suggests that CGG expansions in *NOTCH2NLC* result in at least two different diseases, namely NIID and OPDM. Nevertheless, considering the fact that our patients had peripheral and/or central nervous system involvement together with the clinicopathological features of OPDM, which fills the phenotypic gap between the two diseases, they are most likely in a broad phenotypic spectrum of a single neuromyodegenerative disease rather than two distinct diseases (figure 2S). The identification of the intranuclear inclusions in skin biopsy from two OPDM_NOTCH2NLC patients, which is the diagnostic finding of NIID,^19^ further supports this notion.

A recent research reported that patients with the muscle subtype of NIID have longer CGG repeat expansions from 118 to 517 repeats than those with other NIID subtypes.^6^ Indeed, the CGG repeats in our patients ranged from 116 to 674. Interestingly, the patient (patient 2) carrying 674 repeats, albeit with mosaicism, had milder phenotype than the patient (patient 1) carrying 139 repeats who had an early-onset and severe phenotype, suggesting that there was no apparent correlation between the size of the CGG repeats and the clinical symptoms in OPDM_NOTCH2NLC.

Diagnostically, the presence of intranuclear inclusions stained with anti-poly-ubiquitinated protein, anti-SUMO1, and anti-p62 antibodies in the skin and other organs is pathognomonic of NIID.^19^ In the present study, we confirmed the presence of essentially the same type of intranuclear inclusions in muscle, further suggesting that OPDM and NIID may well be in the same spectrum with the identical degenerative process albeit different organs, such as skeletal muscle, skin, and CNS, are affected in variable degrees. Interestingly, the size of the intranuclear filaments was 12.6 nm in diameter on average on EM, which is almost similar to that observed in neuronal cells in patients with NIID (8–16 nm), ^20 21^ suggesting that the underlying mechanism of the myodegeneration in OPDM_NOTCH2NLC and the neurodegeneration in NIID may well be identical.

In terms of the pattern of muscle involvement on muscle imaging, one study described predominantly involved soleus and long head of the biceps femoris, relatively preserved rectus femoris, and asymmetric involvement pattern in OPDM (without genetic diagnosis),^22^ and another reported markedly involved soleus in patients with OPDM_GIPC1.^4^ The findings of muscle CT in the present three patients with OPDM_NOTCH2NLC are similar to the previous reports,^4 22^ suggesting that the myodegeneration mechanism might be similar between those three OPDM types.

In conclusion, our seven Japanese patients with OPDM_NOTCH2NLC exhibited distinct clinicopathological features, including the involvement of central and peripheral nervous systems. Our findings widen the phenotypic spectrum of a neuromyodegenerative disease caused due to CGG repeat expansions in *NOTCH2NLC*.

## ACKNOWLEDGMENTS

We thank the patients and their families whose help and participation made this work possible. We also thank Dr Noriko Sato for evaluation of MRI. The authors thank Ms. Kaoru Tatezawa, Ms. Naho Fushimi, Ms. Kazu Iwasawa, Ms. Yoko Tsutsumi, and Ms. Kanae Kanna in NCNP for their technical assistance.

## CONTRIBUTIONS

MO, TK, and IN designed studies; AI carried out genetic analysis and analysis of data; ZW contributed to PCR methods; MO, HN, SH, SN, AO and IN evaluated muscle pathology; YO, HK, AN, RA, HT, RH, HD, and FT contributed to the clinical diagnosis and muscle biopsy of patients with OPDM; MO, AI, SH, SN and IN wrote and edited the manuscript; and IN supervised the project; All authors read and approved the final manuscript.

